# Single-cell multi-omic integration analysis prioritizes druggable genes and reveals cell-type-specific causal effects in glioblastomagenesis

**DOI:** 10.1101/2025.06.28.25330486

**Authors:** Yu-Feng Huang, Kun-Long Huang

**Affiliations:** The First Clinical Medical School, Shanxi Medical University, Taiyuan, China

**Keywords:** genome-wide association studies, single-cell multi-omics, glioblastomagenesis, cell-type-specific causal genes

## Abstract

**Background:** Gliomas constitute 80% of malignant brain tumors, with glioblastoma (GBM) being the most aggressive subtype. The single-cell-level mechanisms underlying gliomagenesis are poorly understood, hindering therapeutic development. We combine genome-wide association studies (GWAS) with bulk tissue and single-cell multi-omics to prioritize gliomagenesis genetically supported candidate genes and reveal cell-type-specific biological mechanisms.

**Methods:** We integrated the largest glioma GWAS with brain-specific multi-omics to prioritize genetically supported candidate genes using two broad categories of prioritized methods. Biological enrichment, differential gene expression, and CRISPR/miRNA were used to assess target enrichment and druggability. By integrating single-cell multi-omics data (genomics, transcriptomics, epigenomics), we investigated GBM-relevant cells, tumor microenvironment (TME) interactions, and cell-type-specific mechanisms in glioblastomagenesis. Additionally, phenome-wide association studies (PheWAS) and drug repurposing analyses were conducted to annotate genetic pleiotropy and enhance drug repositioning.

**Results:** We prioritized 11 high-confidence and 47 putatively causal genes, most of which are druggable. Astrocytes and oligodendrocyte precursor cells (OPCs) were identified as the trait-relevant populations in GBM, with significantly increased TME cell communication between these populations and neurons. Fourteen cell-type-specific causal effects in glioblastomagenesis were discovered, including three high-confidence genes (EGFR in astrocytes, CDKN2A in OPCs, and JAK1 in excitatory neurons). Most effects (85.7%, 12/14) were associated with non-GBM-relevant cell cells, encompassing both glial and neural cells.

**Conclusions:** This study systematically identifies genetically supported candidate genes in gliomagenesis and their cell-type-specific effects, providing insights into the cell-resolved mechanisms of glioma susceptibility and advancing targeted precision therapeutics.

## Introduction

Gliomas constitute nearly 30% of primary brain tumors and 80% of malignant cases, with glioblastoma (GBM) being the most prevalent and aggressive subtype, generally associated with a poor prognosis^1^. Despite extensive research, numerous experimental drugs have failed in clinical trials, highlighting the need for more precise and effective therapies^2,3^. However, a major barrier is the intratumoral heterogeneity of GBM^2,4^, resulting from different cells of origin, diverse genetic and epigenetic mutations, and varied tumor microenvironments (TMEs) shaped by cell communication^5–7^. Hence, dissecting cell-type-specific mutations and molecular expression in both cancerous and non-cancerous cells is critical for deciphering gliomagenesis and developing precise interventions^5,8^.

Human genetic evidence markedly improves the success rate of targeted therapies^9^, making large-scale glioma genome-wide association studies (GWAS) and subsequent analyses a powerful approach for understanding gliomagenesis mechanisms and discovering druggable molecules^10–12^. However, single-data approaches often show limited power and explain only a fraction of heritability^13,14^; lack of replication across genetic studies has led to high false-positive rates^15^; and bulk-tissue analyses fail to capture the cell-type-resolved biological processes and cell-type-specific genes^12^, which are linked to higher clinical success^8^.

Because most GWAS-identified risk variants lie in non-coding regions, integrating multi-omics data provides deeper insights for post-GWAS analyses. Protein quantitative trait loci (pQTL) and expression quantitative trait loci (eQTL) datasets offer complementary effects in target investigation^16^. Open chromatin regions and enhancer loops from brain-specific epigenomics data—including ATAC-seq, promoter capture Hi-C (pcHi-C), and the activity-by-contact (ABC) model—improve long-range interactional causal gene mapping^17–19^. Advances in single-cell multi-omics further extend our perspective to single-cell resolution. Large-scale single-cell and single-nucleus RNA sequencing (sc/snRNA-seq) capture global transcriptional profiles at the individual-cell level, thereby improving our understanding of cell-type differentiation and interactions^17,20^. Moreover, cell-type-specific ATAC-seq, ChIP-seq, proximity ligation-assisted ChIP-seq (PLAC-seq), and single-cell eQTL (sc-eQTL) explore cell-type-resolved regulatory consequences and help explain the missing heritability observed in bulk tissue analyses^17,21^.

In this study, we integrate glioma GWAS with brain-specific multi-omics data—including transcriptomics, epigenomics, and proteomics—to identify genetically supported candidate genes for gliomagenesis. By evaluating druggability and target pleiotropy via sc/snRNA-seq, bulk RNA-seq, CRISPR/RNAi, and cross-phenotype GWAS, we prioritized many druggable genes. Single-cell multi-omics were further employed to elucidate cells of origin, TME interactions, and cell-type-specific effects in glioblastomagenesis. **Fig. 1** provides an overview of the main analytical steps conducted in the study.

**Fig 1.**
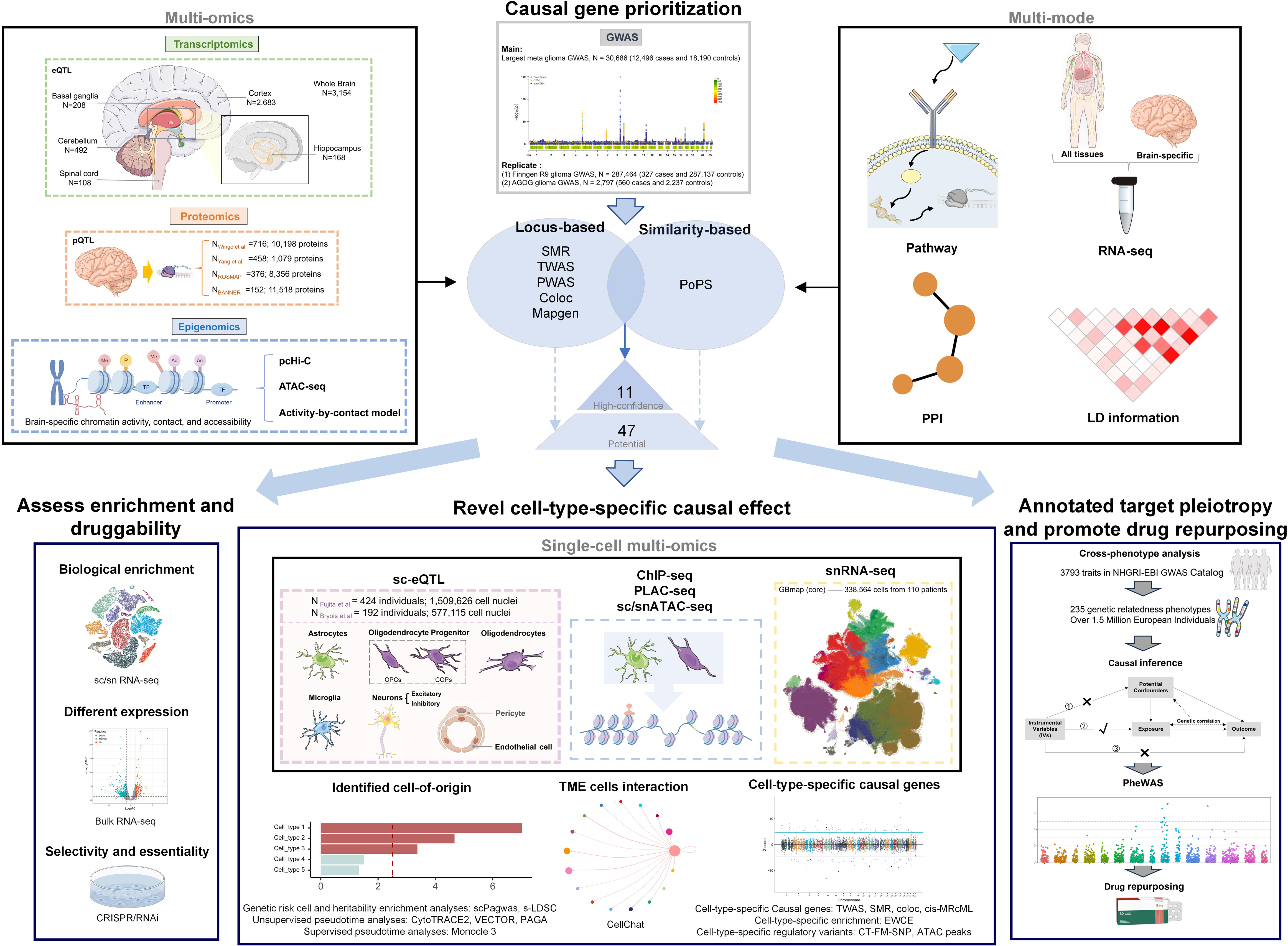
General study workflow. The workflow consists of two main phases: (Top) Candidate gene prioritization: Integration of large-scale GWAS with multi-omics (transcriptomics, proteomics, epigenomics) and multi-mode features to identify high-confidence candidate genes. (Bottom) Downstream multi-dimensional evaluation: Assessing biological enrichment and druggability (left), revealing cell-type-specific causal effects and microenvironment interactions via single-cell multi-omics (center), and annotating target pleiotropy to promote drug repurposing (right).

**Fig 2.**
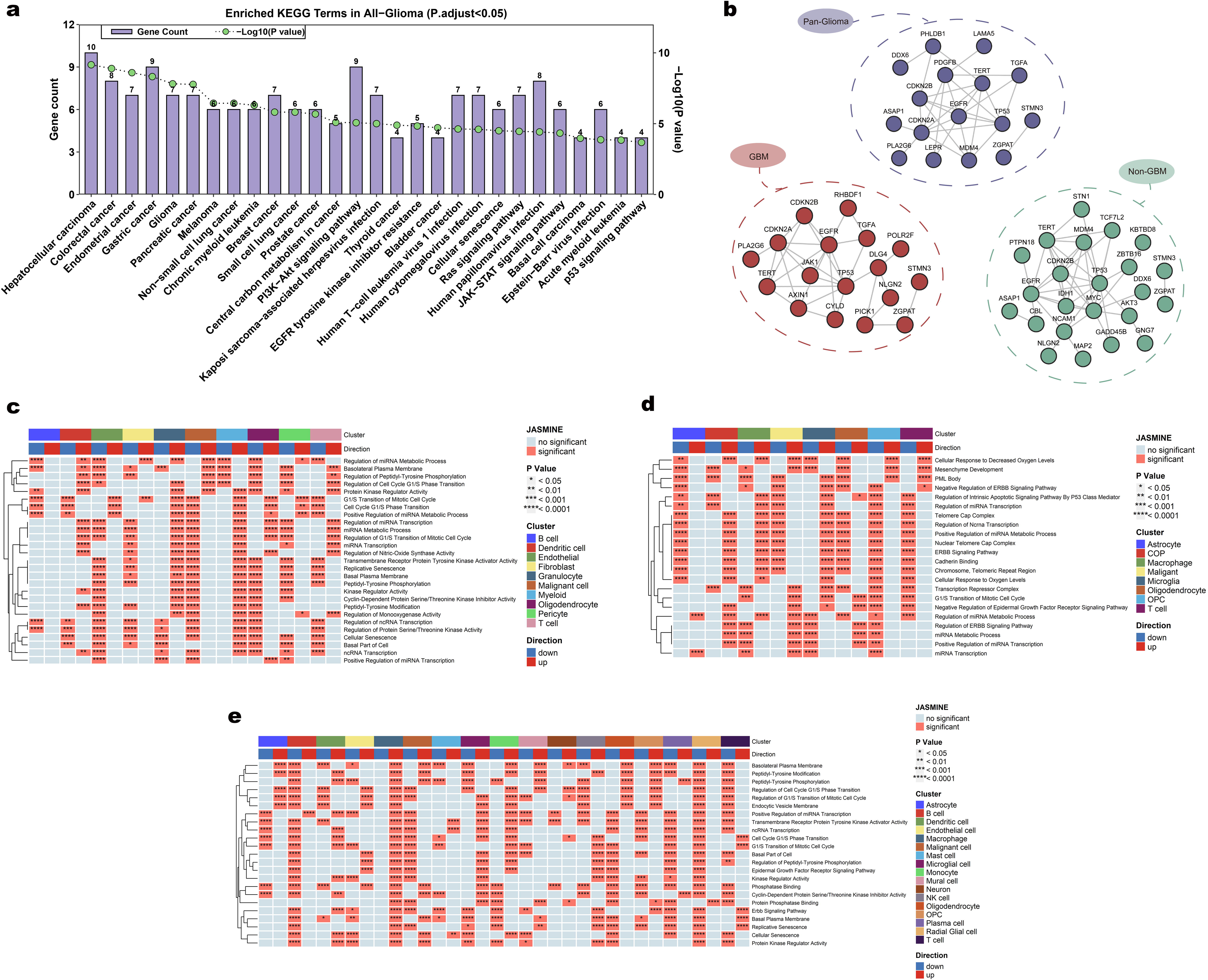
Enrichment of identified genes. **(a)** The bar plot represents the number of genes associated with each KEGG pathway (left Y-axis), while the green dots indicate the -log10(P value) for pathway enrichment (right Y-axis). **(b)** Protein–protein interaction network for identified potential causal genes. **(c-e)** Heatmap of significant gene set distributions across cell clusters. Asterisks indicate P-values: * <0.05, ** <0.01, *** <0.001, **** <0.0001. The left dendrogram shows clustering based on gene set similarity. “Direction” indicates upregulation (red) or downregulation (blue) of gene sets in each cluster. Cell clusters are color-coded as shown.

## Result

### Integrating Brain Tissue-Specific Genetic Multi-Omics Data to Identify Putatively Causal Genes for Glioma

We systematically explored glioma genetically supported candidate genes by integrating large-scale GWAS with brain-specific multi-omics. Two main strategies were employed: a similarity-based method (combining gene expression, pathways, and predicted PPIs) and a locus-based method (incorporating LD data, open chromatin, enhancer loops, and QTL data). Previously reported and newly explored gliomagenesis genes identified by each method are listed in **Supplementary Tables 6, 8–10, 12–14, and 16–18.** Overall, we detected 24, 33, and 26 potential putatively causal genes for pan-glioma, non-GBM, and GBM, respectively **(Supplementary Table 7,11 and 15)**. There were 41 novel glioma-associated molecules that we prioritized. Cross-referencing two broad methods^14^ yielded high-confidence genetically supported candidate genes: 6 in pan-glioma (*EGFR*, *TERT*, *MDM4*, *SOX8*, *CDKN2A*, *STMN3*), 5 in GBM (*EGFR*, *CDKN2A*, *JAK1*, *TERT*, *STMN3*), and 4 in non-GBM (*CDKN2B*, *IDH1*, *NPAS3*, *ZGPAT*). No high-confidence genes were shared by subtypes. Identified putatively causal genes showed a significant enrichment in the cerebral cortex (FC = 2.74, P adj = 3.96E-2), indicating higher drug-target potential due to tissue-specific effects^22,23^. Pathway analyses revealed significant enrichment in cancer-related biological processes (**Fig.2a**), while PPI analysis indicated significant associations among the identified protein-coding gene sets (pan-glioma, P_enrichment_ = 1.54E-04; GBM, P_enrichment_ = 7.86E-04; non-GBM, P_enrichment_ = 1.18E-03) **(Fig.2b)**.

### Verification of Biological Enrichment and Targetability

We assessed the targetability of the identified genes through glioma sc/snRNA-seq, bulk RNA-seq, and CRISPR/RNAi data. First, to evaluate biological enrichment in tumor, we analyzed three large-scale sc/snRNA-seq datasets, scoring GO term enrichment for the identified genetically supported candidate genes. We observed that the target-set-mediated pathways were significantly enriched in both malignant and non-cancerous cells (**Fig.2c-e**), suggesting broad involvement of these genetic target-mediated pathways in glioma^24^.

Next, we matched targets to DEGs (healthy vs. glioma) in five bulk RNA datasets (N total = 1,890). In pan-glioma, GBM, and non-GBM, 62.5–92.3% of identified genes were DEGs in ≥1 dataset, with 18.8–46.2% labeled as DEGs in ≥3 datasets with consistent effect directions. These findings suggest that the identified genes showed significant expression changes between tumor and normal tissues.

Since DEGs may mirror disease-driven expression changes rather than causality^25^, we verified target selectivity and essentiality via genome-wide CRISPR loss-of-function and RNAi viability screens across over 1,300 tumor cell lines. In pan-glioma and its subtypes, the majority of identified genes (88.5–93.8%) were enriched in multiple tumor cell types. Among 72 glioma cell lines, 51.72% (30/58) of targets were selective or essential. Finally, we annotated the identified genes with ligandability assessments, 3D structural data, and compound information. (**Supplementary Tables 7, 11, and 15**)

### Uncovering the Cellular Context Critical For GBM Origins

To determine which cell types contribute to glioblastomagenesis, we combined GWAS data with large-scale GBM snRNA-seq^20^. Malignant cells (P adj = 1.03E-09), astrocytes (P adj = 1.14E-11), and oligodendrocyte precursor cells (OPCs, P adj = 1.51E-10) showed significant associations with GBM risk **(Fig.3b)**. Malignant cells in a proliferative state demonstrated strong differentiation capacities and elevated genetically associated pathway activity scores (gPAS), reflecting high enrichment of cell-specific pathway-based genetic variances^26^**(Fig.3c and 3d)**. Using adult-brain scATAC-seq, we assessed the cell-type-specific heritability enrichment of GBM in trait-relevant cells. OPCs displayed significant GBM heritability enrichment (44.10-fold; P_Enrichment_ = 0.038, P_Coefficient_ = 0.042), while astrocytes did not reach significance (31.97-fold; P_Enrichment_ = 0.075, P_Coefficient_ = 0.082). Pseudotime analyses, derived from GBM snRNA-seq, were consistent with the genetic findings **(Fig 3d-f)**.

**Fig 3.**
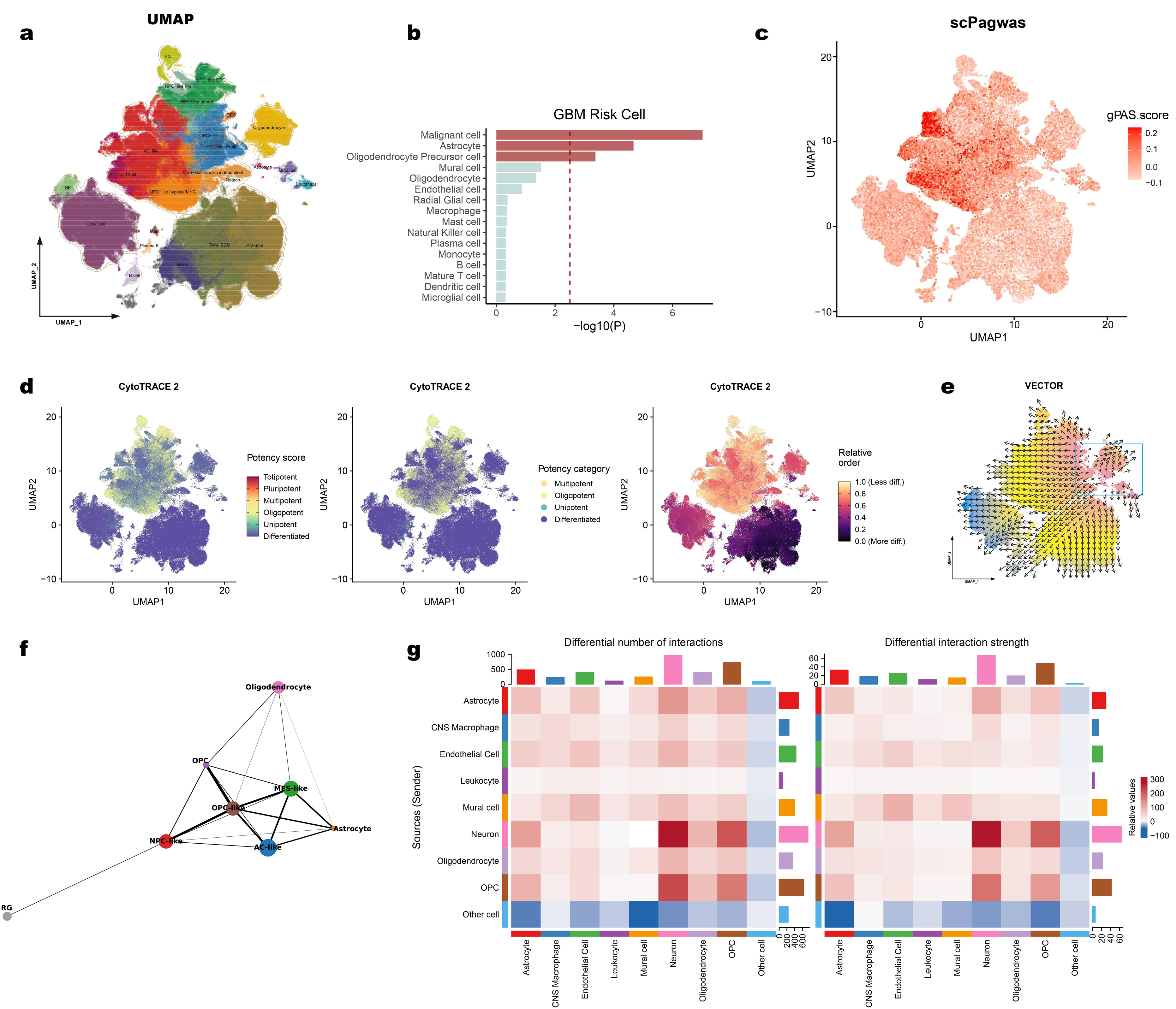
Cell-of-origin and TME cell interactions in GBM. **(a)** UMAP visualization of GBmap (core) **(b)** Bar plot showing genetic GBM-relevant cell types identified in scPagwas, which ranked by p value. Dashed red line indicates the threshold after multiple testing correction. **(c)** UMAP visualization of gPAS scores derived from scPagwas analysis. Cells are colored according to their gPAS score, ranging from low (white) to high (red), indicating regions with higher pathway activity. **(d)** UMAP visualization of differentiation ability colored by CytoTRACE 2 predictions. (Left) UMAP colored by predicted potency score, with values ranging from totipotent (red) to differentiated (blue). (Center) UMAP colored by predicted potency category. (Right) UMAP colored by predicted relative differentiation order, transitioning from less differentiated (yellow) to more differentiated (purple). **(e)** UMAP visualization of pseudotime by VECTOR, the pseudotime order of cells is depicted by color, transitioning from red to blue. The blue rectangle highlights the region corresponding to the starting cells. Vector representation of inferred developmental directions for cells in GBM. **(f)** Graph plot illustrating differentiate relationship identified by PAGA, Nodes represent cell types, and edge thickness reflects connectivity strength. **(g)** Heatmaps showing the differences in brain cell interactions between GBM non-malignant cells and healthy controls in GBM high-incidence brain regions. interaction number (left) and interaction strength (right) among cell populations across two datasets. Rows represent source (sender) cell types, and columns represent target cell types. The top bar plot shows the total incoming signaling for each target cell type, while the side bar plot indicates outgoing signaling for each source cell type. Red indicates increased signaling in the GBM patient dataset compared to the healthy dataset, and blue indicates decreased.

### Cell Interaction in the GBM TME

To investigate cell interactions in the GBM TME, we compared cell-cell communication among non-cancerous cells from patients and healthy controls. In total, 1,129,369 cells and 846 signaling genes were included. We observed significantly elevated cell-cell communication in GBM **(Supplementary Fig. 4)**, especially between neurons and trait-relevant cells (astrocytes, OPCs). **(Fig 3g)**.

### Investigating Cell-Type-Specific Effects

We performed cell-type-specific causal gene analyses based on sc-eQTL data from eight brain cell types. 14 cell-type-gene pairs involving 12 genes were identified, with 58.3% being novel in brain tissue analyses. Three high-confidence candidate genes—*JAK1*, *CDKN2A*, *EGFR*—showed cell-type-specific effects **(Fig 4a-e and Supplementary Fig. 6)**. In GBM-relevant cells, we detected cell-type-specific glioblastomagenesis effects, including *EGFR* in astrocytes and *CDKN2A* in OPCs. The majority of detected cell-type-specific effects (85.7%, 12/14) involved non-GBM-relevant cells, including both glial and neural cells. Oligodendrocytes accounted for most of the cell-type-specific genes (8/12). Excitatory neurons were enriched in high-neural GBM, and their neural-tumor interactions drive glioma growth^27^; their cell-type-specific causal gene *JAK1* was also a high-confidence causal gene in tissue-level analyses.

**Fig 4.**
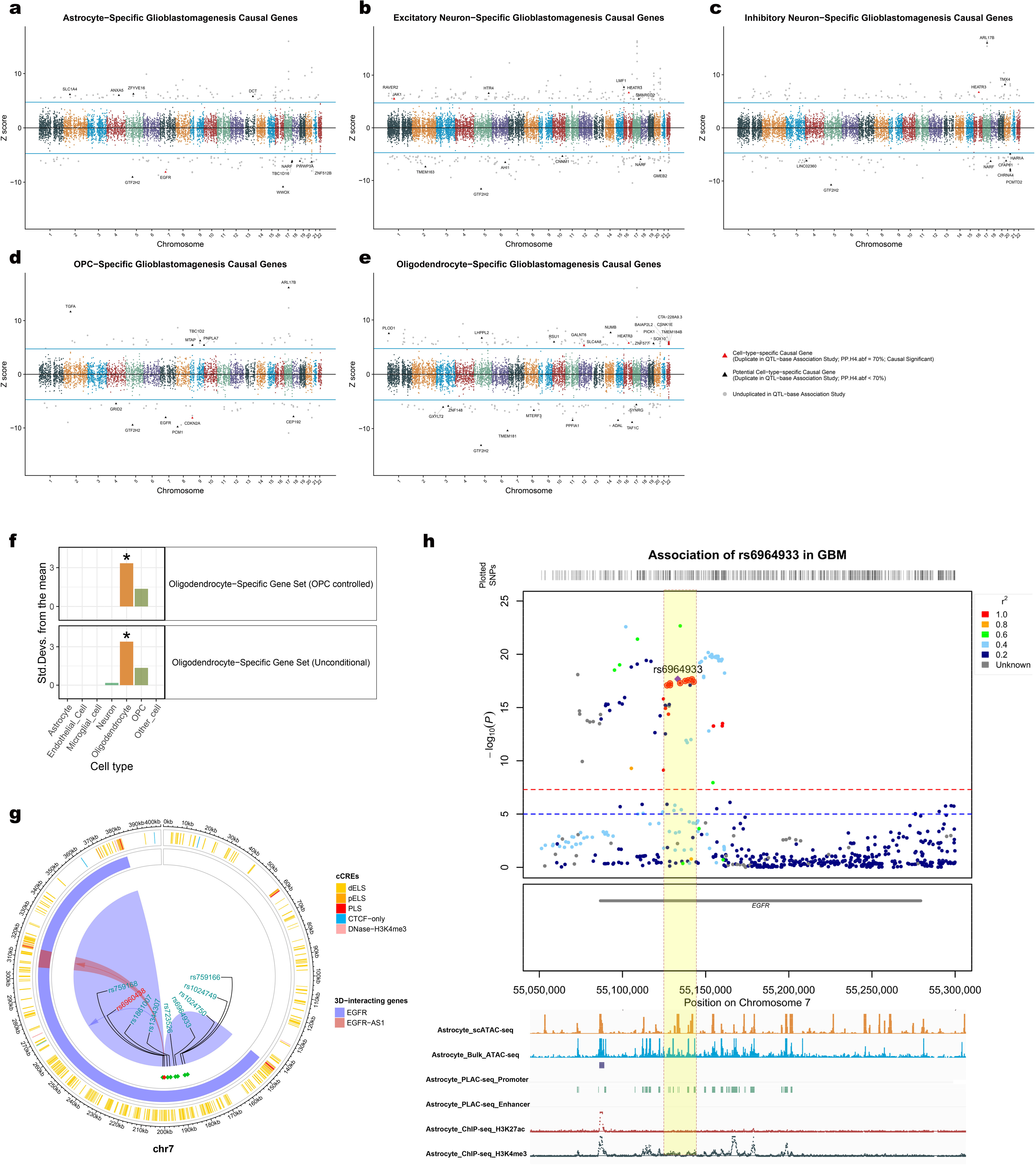
Cell-type-specific causal effect in glioblastomagenesis. (a-e) Manhattan-style plots displaying cell-type-specific glioblastomagenesis causal genes across chromosomes for astrocytes, excitatory neurons, inhibitory neurons, OPCs, and oligodendrocytes. Z-scores represent gene effect size and direction in QTL-based association study, with significant cell-type-specific causal genes highlighted as red triangles (Duplicated in QTL-based association study, PP.H4.abf ≥ 70%, and with causal significance) and potential causal genes as black triangles (Duplicated in QTL-based association study, and PP.H4.abf < 70%). Grey dots indicate unduplicated genes from QTL-based association studies. Horizontal blue lines denote significant thresholds (P= 2.78e-6; 0.05/17,964). **(f)** Bar plots showing the cell type enrichment test in significant oligodendrocyte-specific gene sets. (Top) Results for OPC-controlled analysis. (Bottom) Results for unconditional analysis. Asterisks (*) highlight significant deviations, indicating overexpression in oligodendrocytes compared to the bootstrapped mean. **(g)** Circular plot illustrating cis-regulatory elements (cCREs) and 3D genomic interactions in rs6964933 and its high LD credibleset. Outer tracks show cCREs, categorized as dELS (yellow), pELS (orange), PLS (red), CTCF-only (blue), and DNase-H3K4me3 (cyan). Inner arcs represent 3D interactions between regulated genes. SNPs of interest are labeled and linked to their associated regions. Purple and red shading indicates chromatin loops connecting regulatory elements to target genes. Visualization by the ViSNP package. **(h)** Regional Manhattan plot showing rs6964933 and its high LD credible set in GBM GWAS. The y-axis displays -log10(p-values), and points are colored based on linkage disequilibrium (LD) with rs6964933. Horizontal dashed lines indicate genome-wide significance (5×10^-8) and suggestive significance (1×10^-5) thresholds. Peak plot generated by IGV v2.17.2 displayed with additional tracks for astrocyte-specific epigenomic peaks, including snATAC-seq, bulk ATAC-seq, regulatory elements from PLAC-seq, and histone modifications (H3K27ac and H3K4me3) from ChIP-seq. The highlighted yellow region marks the position of rs6964933 and its high-LD variants credible set.

To evaluate cell-type-specific enrichment of identified genes, we performed conditional/unconditional enrichment analyses on large-scale GBM snRNA-seq. Among the significant cell-type-specific gene sets, only oligodendrocytes had ≥4 genes for further analysis. We found that oligodendrocyte-specific targets exhibited high specificity in oligodendrocytes (P adj = 3.92E-2). Even after controlling for OPCs, oligodendrocyte enrichment remained significant, suggesting independence from OPC enrichment. To further probe enrichment in other cell types, we evaluated cell-type-specific gene groups (significant in association studies and replicated). Each of the four cell-type-specific gene groups (neuron, astrocyte, OPC, oligodendrocyte) displayed maximal specific expression in their respective cell types; two (neuron, oligodendrocyte) reached significance (<0.05) following Bonferroni correction **(Supplementary Fig. 6)**.

We integrated GBM GWAS fine-mapping results with over 900 cell-type-specific cCREs to uncover cell-type-specific causal variants in glioblastomagenesis. We identified rs6964933, located within the *EGFR* gene body, as an astrocyte-specific causal variant (PIP = 0.10, Z-score = 1.99). This variant also appeared in the GWAS fine-mapping credible model set (P_GWAS_ = 2.19E-18, PIP = 0.18) **(Supplementary Table 20)**. Within the rs6964933 high LD SNP set identified by fine-mapping (N_SNP_= 9, R² > 0.95, PIP_sum_ = 1), we detected high FORGEdb scores (range 6 to 10), and many SNPs overlapped astrocyte-specific epigenomic peaks, suggesting a potential effect on cell-type-specific transcriptional regulation^28,29^ **(Fig. 4g-h)**.

### High *EGFR* Expression Related to Lower Genetic Glioblastomagenesis Risk

While somatic EGFR alterations are well-established oncogenic drivers in formed GBM^3031^, our genetics and QTL framework reveals a distinct, cell-context specific role for EGFR in germline susceptibility. Separating this from somatic oncogenesis, our eQTL/pQTL analyses demonstrate an inverse association between genetically predicted EGFR expression (mRNA/protein) and GBM susceptibility, an effect predominantly driven by astrocyte-specific regulatory variants **(Fig. 5a) (Supplementary Tables 11 and 19)**. Robust colocalization of QTL and GWAS signals further supported this, mediated by shared germline variants that exhibit opposing directional effects on astrocytic EGFR levels and overall GBM risk **(Fig. 5b-f)**. To contextualize this baseline genetic association within disease progression, we performed supervised pseudotime analyses using normal astrocytes as root cells. EGFR expression varied dynamically across the predicted differentiation trajectory from astrocytes into malignant, AC-like cells **(Fig. 5g-h)**. Furthermore, an exploratory comparison between normal brain cells and GBM samples suggested a potential upregulation of EGF pathway interactions (including EGFR and its ligands) within the tumor microenvironment (TME), particularly in associated astrocytes **(Fig. 5i and Supplementary Fig. 7)**. Together, these findings indicate that while genetically determined EGFR expression in normal astrocytes is inversely associated with GBM susceptibility, distinct constitutive and ligand-driven somatic EGFR signaling dynamics emerge later during glioblastomagenesis and TME remodeling^32^.

**Fig 5.**
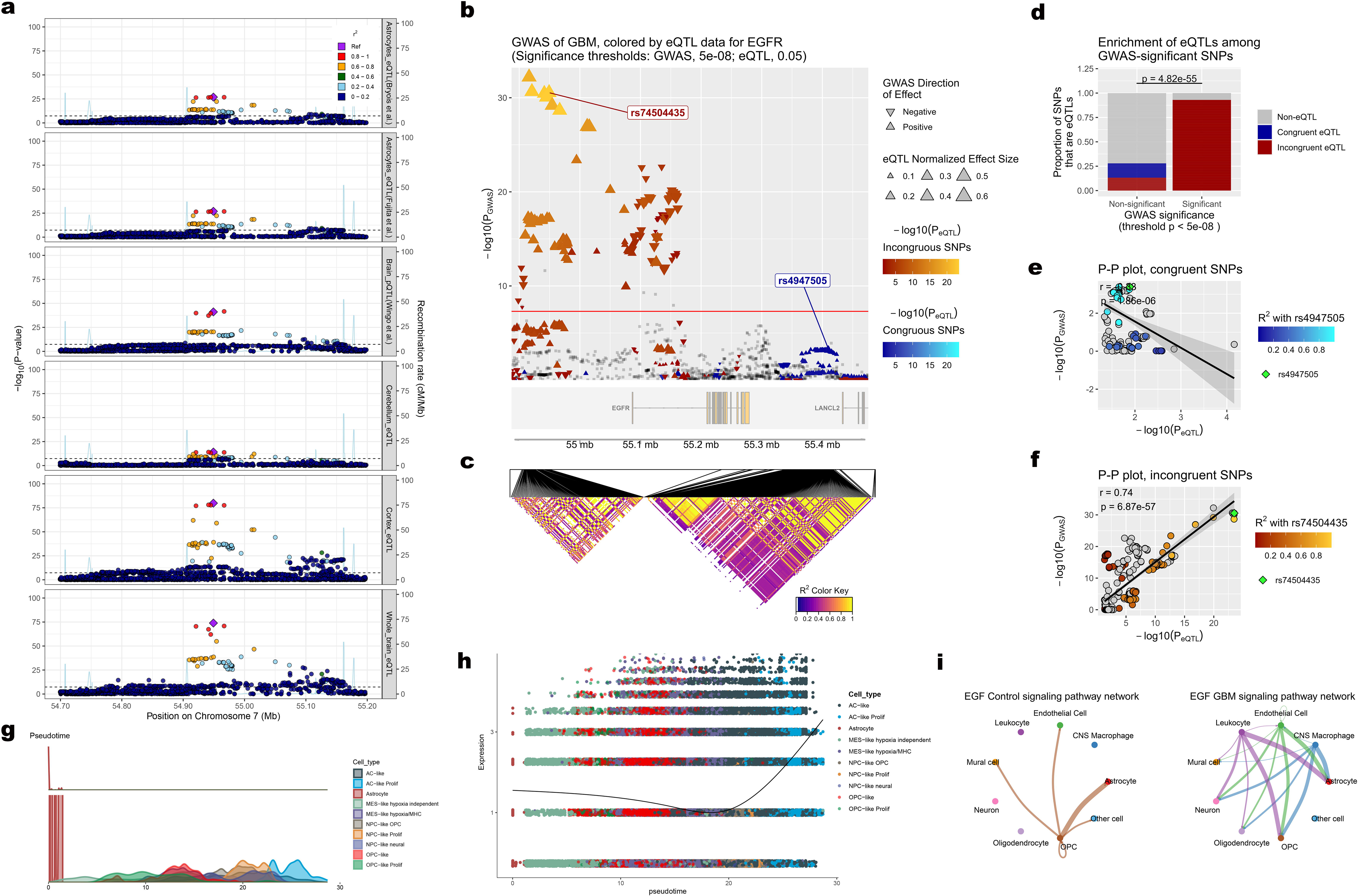
EGFR effect in glioblastomagenesis. (a) Regional association plots for distinct brain QTL datasets showing genetic signals in the EGFR cis region. Points represent SNPs, colored by LD (R2) with the lead SNP (purple diamond). The y-axis displays -log10(p-values) for associations, and the light blue line indicates recombination rates across the region. The dashed horizontal line denotes the significance threshold (5×10^-8). (b) eQTpLot visualization of the colocalization and driving effect between GBM GWAS and astrocyte-specific cis-eQTL data (Fujita et al.) in the EGFR cis region. Manhattan plot showing GWAS signal, with SNPs colored by eQTL P value and direction (triangles for positive and negative effects). Genomic region plot showing the genomic structure. (c) LD heatmap depicting pairwise LD across the locus. (d) Bar plot quantifying the enrichment of eQTLs among GWAS-significant SNPs (p<5e−8), distinguishing congruent and incongruent eQTLs. (e) P-P plot correlating GWAS and eQTL p-values for congruent SNPs, (f) P-P plot correlating GWAS and eQTL p-values for incongruent SNPs.(g) Density plot demonstrates cell type distributions along pseudotime inferred by Monocle3, with astrocytes designated as the starting cell population. Each curve represents the relative proportion of a specific cell type as pseudotime progresses, highlighting differentiation trajectories.(h) Pseudotime trajectory plot showing expression dynamics of EGFR along pseudotime, with astrocytes set as the starting cell population in Monocle3. Points represent individual cells, colored by cell type, and the black curve indicates the smoothed trend of EGFR expression across pseudotime. (i) Circle plot comparison of EGF signaling pathway networks between healthy controls and GBM non-malignant cells. The edge color represents the source cell group as the sender, and the edge weight reflects interaction strength, with thicker lines indicating stronger signals.

### Exploring Causal Phenotypes and Target Gene Pleiotropy

Safety concerns and side effects frequently terminate oncology clinical trials^23^, yet glioma risk factors remain unclear. To explore glioma causal phenotypes and genetic pleiotropy of target genes, we undertook several analyses. First, we examined the genetic architecture of 3,000 phenotypes, identifying 137 (pan-glioma), 67 (GBM), and 130 (non-GBM) significantly associated phenotypes (P adj < 0.05). GWAS summary data were collected from 260 phenotypes (126 pan-glioma, 105 GBM, 152 non-GBM). Using a two-sample MR framework, we performed a Mendelian randomization phenome-wide association study (MR-PheWAS) **(Supplementary Fig. 8)** After heterogeneity, pleiotropy, and MR sensitivity assessments, we found 7, 7, and 9 potential causal traits for pan-glioma, GBM, and non-GBM, respectively. In light of widespread genetic heritability, we applied the LCV analysis on potential genetically correlated (P<0.05) glioma-phenotype pairs^33^. There were 5, 6, and 5 suggestive causal traits identified for pan-glioma, GBM, and non-GBM, respectively **(Supplementary Tables 22)**. Only telomere length showed robust causal evidence for pan-glioma after Bonferroni correction (OR = 1.888, 95% CI (1.521 to 2.344), P_IVW_= 8.37E-9, P_HDL_ = 3.31E-1). Notable trends emerged among other potential traits across various data. For instance, traits reflecting cognitive proficiency (e.g., good cognitive function, high educational attainment, high intelligence) correlated with reduced non-GBM risk. Conversely, schizophrenia, a disease characterized by cognitive impairment, aligned with a higher non-GBM risk. Finally, we conducted a gene-based PheWAS to characterize target pleiotropy^34^. Reported pleiotropy mostly concerned potential causal phenotypes. **(Supplementary Tables 23)**.

### Drug Repurposing of Target Genes

To facilitate drug repurposing for identified targets, we queried databases to detect drugs with high gene-specific interaction scores (>0.9). We then annotated drug characteristics along with the latest clinical details. We collected 87 drugs targeting 28 genes in more than one clinical trial: 57 (65.51%) for tumor therapy, 21 (24.13%) for brain diseases, and 18 (20.69%) for glioma therapy. **(Supplementary Tables 24)**

## Discussion

In this study, we integrated large-scale GWAS with brain-specific multi-omics (transcriptomic, proteomic, and epigenomic) data and employed two broad categories of gene prioritization strategies to investigate the genetically supported candidate genes for gliomagenesis. 47 potential and 11 high-confidence putatively causal genes were identified. Evidence from large-scale sc/snRNA, bulk RNA, and CRISPR/miRNA data showed that these targets were significantly enriched in glioma pathways, differentially expressed (patients vs. controls), and selective or essential in cancer cell lines. To elucidate cell-type-specific effects, we integrated single-cell multi-omics data with GBM GWAS, enabling us to pinpoint cells of origin, examine TME cell interactions, and detect multiple cell-type-specific causal genes. We identified 12 causal genes mediated by cell-type-specific effects in risk and other non-malignant cells (both glial and neural cells), potentially linked to elevated cell interactions in the GBM TME. This underscores the importance of cell-type-level precision therapeutics. To refine target pleiotropy assessment and advance drug repositioning, we conducted PheWAS and drug repurposing analyses.

Compared to previous tissue-level studies^10–12,35–39^, we prioritized 41 novel glioma-associated molecules, Notably, two of these genes were in genome-wide significant loci that had not been linked to any genes^10^. All previous glioma gene prioritization studies using locus-based methods have especially focused on the QTL-based method, which may relate to lower precision (14–46%) due to a fraction of heritability explained by QTLs and static power limitations^13^. We combined two different methods and highlighted 11 high-confidence putatively causal targets for gliomagenesis, which provide a precision of at least 67% and up to 79% in previous benchmarked analyses. Notably, no high-confidence targets overlapped between GBM and non-GBM, reflecting glioma subtype heterogeneity^7^.

GBM heterogeneity correlates with multiple cells of origin^1,7,40^. Here, astrocytes and OPCs emerged as GBM origin cells, and related cell-type-specific causal genes expand our understanding of glioblastomagenesis. *CDKN2A* is deleted in approximately 60% of GBM, sensitizing malignant cells to lipid peroxidation and ferroptosis^41^. Recent work demonstrates that methylation-silenced *CDKN2A*, along with OPC insulator loss, further drives gliomagenesis^42^. Here, we determined its OPC-specific causal effect on glioblastomagenesis. *EGFR* is one of the most promising GBM therapy targets^3^, amplified in ∼50% of GBM patients^43^. Recent study indicates EGFR-amplified malignancies tend to differentiate into astrocytic lineages, and in some cases, *EGFR* amplification is not involved in early glioblastomagenesis^44^. Here, we pinpointed an astrocyte-specific causal effect of *EGFR* and its variant rs6964933 in glioblastomagenesis. Interestingly, increased astrocyte-specific *EGFR* expression was associated with lower GBM risk, possibly associated with *EGFR* ligand activity across malignant transformation and TME development stages^31,32^.

The majority of detected cell-type-specific effects (85.7%, 12/14) involved non-GBM-relevant cells, including both glial and neural cells. Oligodendrocytes, which are abundant and drive neuronal activity in the GBM TME^27^, accounted for 66.7% cell-type-specific genes. Recent spatial and single-cell transcriptomic analyses reveal that mature oligodendrocytes are not merely passive structural components but actively shape a pro-tumorigenic TME. Under tumor-induced stress, mature oligodendrocytes can adopt a reactive state, secreting supportive cytokines and chemokines (such as CCL5) that interact with receptors on glioma stem cells to maintain tumor stemness and drive treatment resistance^45^. Furthermore, their dysregulation and aberrant signaling during neuroinflammation can create a permissive niche that facilitates early tumor initiation^45^. Neurons constitute the largest brain cell population, and intratumoral neurons often exhibit high excitability, contributing to glioma development and progression^27^. Functionally, active neurons drive glioma progression through both paracrine signaling and direct electrochemical integration. Neurons release neuromodulatory factors, including brain-derived neurotrophic factor (BDNF) and neuroligin-3 (NLGN3), which activate oncogenic signaling pathways (e.g., PI3K/mTOR) in adjacent glioma cells. Crucially, recent breakthroughs have demonstrated that glioma cells form bona fide AMPA receptor-dependent glutamatergic synapses with neurons. This synaptic integration creates a hyperexcitable network that synchronizes tumor cell proliferation, accelerates invasion, and fosters an immunosuppressive TME^46,47^.By comparing non-malignant TME cells with healthy controls, we found significantly elevated communication—particularly between neurons, astrocytes, and OPCs—likely linked to neuronal hijacking in tumorigenesis^48^. Intratumoral neuronal expression also affects GBM immunotherapy efficacy^49^. Here, we identified *JAK1*, a component central to many pro-inflammatory cytokine signaling and its neuronal expression regulates neuropeptide expression and neuroinflammation across various tissues^50^, as an excitatory-neuron-specific causal gene for glioblastomagenesis.

Many genetic molecules associated with gliomagenesis have been used in glioma therapy or clinical experiments such as *IDH1*, *EGFR*, and *TERT*. To improve the clinical translation, we assessed the targets based on tumor data. The identified target was significantly enriched in glioma biological pathways and differential expression between glioma patients and healthy individuals, most importantly, CRISPR and RNAi screens showed that 53.6% of the novel genes exhibited either selective dependency or essentiality in glioma cell lines. Besides, the robust genetic evidence and brain tissue/cell-type enrichment in our identified molecules are all associated with higher clinical success^8,9,22^. The targets were annotated with chemical structure information and genetic pleiotropy to identify potential side effects^23^. In the context of the central nervous system (CNS), evaluating such pleiotropy is particularly critical, as off-target drug engagements in healthy brain tissues can lead to severe dose-limiting neurotoxicity^51^. Furthermore, successful translational neuro-oncology heavily relies on overcoming the formidable blood-brain barrier (BBB). Therefore, we prioritized actionable targets that are amenable to modulation by small molecules with favorable CNS pharmacokinetic properties^52^.The up-to-date clinical experiments were collected to promote drug repurposing; several promising drugs for glioma were uncovered; for example, Tertomotide hydrochloride, an approved TERT inhibitor for pancreatic cancer^53^, crosses the BBB effectively and demonstrated promising results for intracranial lesions in recent trials^54^. Highlighting such candidates underscores the translational relevance of our pipeline: we can strategically bypass the most significant attrition point in neuro-oncology drug development, thereby accelerating the bench-to-bedside translation for glioma patients^51^.

Despite notable findings, several limitations exist. First, our glioma GWAS subtypes are based on classifications established before the WHO 2021 guidelines, which integrate molecular data, thereby reclassifying IDH-mutant high-grade astrocytoma under the non-GBM group^55,56^. However, earlier studies suggests that potential misclassification did not bias PRS estimates from this GWAS^57^, and we found that the characteristic molecular marker *IDH1* was only identified in the non-GBM data. This may suggest that most GBM and non-GBM cases were likely IDH-wildtype and IDH-mutant, respectively. Given non-GBM subtype heterogeneity^55^, we primarily focused on GBM cell-type-specific genes. Future efforts should refine GWAS using updated glioma subtype definitions. Second, our analyses relied on brain-specific data^58^, and the detected genes show significant enrichment in brain tissues and cells, potentially boosting clinical trial success^22^ while lowering systemic side-effect risks^23^. However, the BBB remains a major hurdle in glioma therapy, impeding drug penetration^2^. In contrast, plasma targets may not require BBB crossing, facilitating delivery. To address this, we assembled current clinical trial data on the identified targets and highlighted drugs tested in CNS diseases. Third, considering the high false positive rate in QTL association study^14^, we conducted replicate studies based on different QTL and GWAS. Although we set a relaxed replication threshold (P=0.05), the disparity in sample sizes between discovery and replication could inflate false negatives. Fourth, regarding our cell-cell communication analyses, although we utilized the largest available GBM single-cell atlas alongside a comprehensive multi-region healthy human brain atlas, comparing cross-dataset scRNA-seq cohorts inherently introduces technical confounders such as variations in sequencing depth, processing protocols, and donor-level heterogeneity. Future single-cell studies profiling paired GBM and adjacent morphologically normal tissues from the same individuals will be crucial to strictly control for these variables and definitively validate these TME signaling networks. Fifth, our analysis mainly focused on European ancestry and many of the glioma germline variants are European specific, which constrains generalizability to other populations. Finally, our study was based on multi-omics evidence from the largest datasets, which is highly reproducible and easy to replicate, but future experimental validation remains essential for clinical translation.

In summary, we applied tissue- and cell-type-specific multi-omics data to systematically dissect genetically supported candidate genes in gliomagenesis, shedding light on cell-type-level mechanisms and advancing future precision medicine efforts.

## Method

### GWAS data

We used the GLIOGENE consortium glioma meta GWAS of 30,686 European (EUR) individuals (6,183 GBM, 5,820 non-GBM, and 18,169 controls) for the main analyses^10^. We included two additional EUR glioma GWAS from FinnGen R9 (243 GBM, 84 non-GBM, 287,137 controls)^59^ and the Australian Genomics and Clinical Outcomes of Glioma (AGOG) (560 glioma, 2,237 controls)^36^ for replicated QTL-based analyses (SMR, PWAS, TWAS). GWAS data for causal inference included 209 phenotypes **(Supplementary Table 1 and 21)**. Additional data and related information are available in the **Supplementary Methods**.

### Gene prioritization using similarity-based methods

#### (1) PoPS

Polygenic Priority Score (PoPS, v0.2) integrates GWAS, gene expression, biological pathways, and predicted protein-protein interactions (PPIs) to prioritize genes^14^. Analyses were performed independently for all-tissue and brain-specific features. We partitioned the genotype into 1,703 LD blocks using LDetect^60^, then we ranked the top 500 (<3%) genes and reported the highest-scoring gene per genome-wide significant LD block (where the number of genome-wide significant SNPs > 1).

### Gene prioritization using locus-based methods

#### (1) Mapgen

We fine-mapped genome-wide significant blocks with L=10 via mapgen (v0.5.8)^61^. We integrated brain-specific epigenomics to capture long-range regulation. The gene-level posterior inclusion probability (PIP) is the weighted sum of PIPs for all linked SNPs. We reported the gene with the highest PIP in genome-wide significant LD block when PIP_gene_ > 0.8.

#### (2) TWAS

We conducted a transcriptome-wide association study (TWAS) using FUSION^62^, which integrates GWAS with precomputed gene expression models^63^ to identify phenotype-related genes. The largest brain-specific cis-eQTL datasets from whole brain (N = 3,154)^64^ and five brain regions (N =108 to 2,683)^65^ were included **(Supplementary Table 2)**. A robust GCTA-GREML (v1.94.2)^63,66^ was used to generate heritability estimates per feature and the likelihood ratio test P value. Only features with P<0.05 and H2>0.1 were retained for analysis. An ACAT-O omnibus test (v0.91)^67^ combined results from all predictive models, generating meta-P values and effect sizes^68^.

#### (3) PWAS

We conducted a proteome-wide association study (PWAS) using four brain-specific pQTL datasets from the EUR population: Wingo et al.^69^, with 10,198 proteins and N = 716; The Religious Orders Study and Rush Memory and Aging Project (ROS/MAP) cohort^70^, with 8,356 proteins and N = 376; The Banner Sun Health Research Institute (Banner) cohort^70^, with 11,518 proteins and N = 152; and Yang et al.^71^, with 1,079 proteins and N = 380 **(Supplementary Table 3)**. This method connects genes and phenotypes through functional protein variation.

#### (4) SMR

We performed summary-level Mendelian randomization (SMR, v1.3.1) analyses^72^ based on previous QTL data. HEIDI tests (P_HEIDI_ >0.05) were applied to distinguish pleiotropy from linkage.

#### (5) Replication studies

We conducted two separate replication studies using different QTLs and GWAS datasets in the QTL-based association analyses (SMR, PWAS, and TWAS): (1) various QTL sources and GLIOGENE glioma GWAS, (2) GWAS datasets from FinnGen and AGOG, along with prior QTL data. The AGOG GWAS was used in the GBM replication study because the majority of its cases were GBM^36^. Gene-disease pairs that satisfied (P_SMR_ <0.05 &P_HEIDI_ >0.05 or P_TWAS/PWAS_ <0.05) in two duplicate studies were considered successfully replicated.

#### (6) Colocalization

Colocalization analyses enhance QTL association study results by excluding genetic confounding^73^. We applied colocalization focusing on the cis gene region using coloc (v5.2.3). Colocalization analysis evaluates five hypotheses regarding the association of the variants with the traits. Posterior probability H4 represents the hypothesis that the variants are associated with both traits and share the same causal variant. Genes displaying significant association (P < 2.12e-6, Bonferroni correction for 23,517 genes) in the QTL-based association analyses, successfully replicated, and demonstrating colocalization evidence (SNP.PP.H4 > 0.70) were reported. Additional detailed information can be found in the **Supplementary Methods**.

### Enrichment Analyses and PPI

We first assessed tissue enrichment of the identified genetically supported candidate gene set via TissueEnrich (v1.24.1)^74^, adjusting P values by Bonferroni correction. PPI analyses utilized the STRING database (v12.0)^75^. The biological functions of the gene set were annotated using the Gene Ontology (GO) and Kyoto Encyclopedia of Genes and Genomes (KEGG) databases through the clusterProfiler^76^. To evaluate pathway activation in tumors, we performed single-cell Gene Set Enrichment Analysis (scGSEA) using the tumor-adapted method JASMINE^77^ on glioma subtype sc/snRNA-seq data. Three sc/snRNA-seq datasets were included: (1) Pan-glioma scRNA-seq containing 55,284 cells from 11 glioma patients^78^ ; (2) GBM snRNA-seq from GBmap (core), containing 338,564 cells from 110 IDH-wild-type GBM patients^20^; and (3) Non-GBM scRNA-seq containing 76,639 cells from 16 IDH-mutant glioma patients^79^. **(Supplementary Table 5)**. Gene sets with significant GO terms (P_Bonferroni_ <0.05) were chosen for scoring.

### DEG

We conducted two differential gene expression (DEG) analyses across five bulk RNA-seq datasets **(Table S4)**. RNA base DEG analysis was carried out, to compare healthy (N_control_ = 1,167) most of from GTEX^80^ with glioma (N_case_ = 1,728) from the CGGA^81^ and TCGA^82^, with provided classifications. Briefly, datasets were normalized with edgeR (v4.0.3) using TMM (trimmed mean of M-values)^83^. Then log2CPM values were computed by voom. Batch effects across datasets were estimated using surrogate variable analysis (SVA) and included as covariates in the limma (v3.58.1) model^84^. Two gene expression microarray datasets were utilized for within-dataset contrasts (normal vs. tumor) in DEG analyses via the online tool GEO2R^85^. Limma precision weights (voom) estimated the mean-variance relationship and produced observational-level weights. A gene was considered differentially expressed when the logFC > 1 and P adj < 0.05.

### DepMap

To evaluate gene selectivity and essentiality for tumor cells, we examined the identified genes in the DepMap (24Q2) dataset (https://depmap.org/portal), which offers genome-wide CRISPR and miRNA loss-of-function screen data from characterized cell lines^86^. Target selectivity and essentiality (gene effect < -0.5) from CRISPR and RNAi screens on 72 glioma cell lines.

### CellChat

To compare cell-cell communication between the GBM and healthy brain, we computed the communication probabilities among non-cancerous cell types in brain regions (frontal, temporal, and parietal lobes) with high GBM incidence^87^ using CellChat (v2.1.2)^88^ and compared them between GBM patients and healthy controls. Two large-scale snRNA-seq datasets were included in: (1) Case: GBmap (expanded), including 289,395 cells from 240 IDH-wild-type GBM patients^20^; and (2) Control: the Human Brain Cell Atlas, containing 839,975 cells from 3 healthy adults^89^**(Supplementary Table 5)**. A custom human ligand-receptor (L-R) database was used, which was generated by merging CellChatDB and CellPhoneDB (v5.0.1)^90^, increasing curated L-R interactions from 3,234 to 6,007 and complexes from 338 to 700.

### Genetic Trait-relevant Cell Analyses

To distinguish critical cell types for glioblastomagenesis, we applied scPagwas (v1.3.0), a pathway-based polygenic regression method integrating GWAS signals with sc/snRNA-seq transcriptional features^26^. The GBM meta GWAS and GBmap (core) snRNA-seq data^20^ were used as inputs; cell types with P_Bonferroni_ <0.05 were deemed GBM-relevant cells. Then, we assessed the GBM-relevant cell-type-specific (astrocytes, OPCs) heritability enrichments (proportion of heritability explained divided by annotation size) and estimated the proportion of the cell-type annotations by applying S-LDSC (v1.0.1)^91^ in adult brain scATAC-seq^92^. The latest version of the baseline model (v2.3)^93^ was used.

### Pseudotime Analyses in snRNA-seq

We implemented a deep learning-based method, CytoTRACE2 (v1.0.0)^94^, to predict cellular potency and developmental potential in GBM. The results were projected onto a UMAP. Unsupervised inference methods VECTOR (v0.0.4)^95^ and PAGA^96^ identified starting cells and inferred developmental trajectories in glioblastomagenesis. We defined astrocytes and OPCs as root cells and inferred their differentiation in GBM using semi-supervised method Monocle 3 (v1.4.18)^97^.

### Cell-Type-Specific Causal Target Genes Analyses

We incorporated two sc-eQTL datasets spanning eight major brain cell types^21,98^ to pinpoint cell-type-specific causal genes in glioblastomagenesis, one by Fujita et al.^98^, involving 1,509,626 cells from 424 EUR individuals, and another by Bryois et al.^21^, involving 577,115 cells from 192 EUR individuals **(Supplementary Table 2)**. Cell-type-specific TWAS and SMR were performed, the genes displaying significant association considered significant (P < 2.78e-6, Bonferroni correction for 17,964 genes). Two replicate studies used independent QTLs and alternative GWAS data. FinnGen and AGOG GWAS replicated our QTL-based association analyses (TWAS, SMR). Genes that replicated (P_TWAS_ <0.05 or P_SMR_ <0.05 and P_HEIDI_ >0.05) were labeled as potential cell-type-specific putatively causal genes. Colocalization analysis (PP.H4 >0.7) was conducted to improve identification of cell-specific effects^61^. cis-MRcML (v0.0.09), which an MR method based on conditional/joint SNP effects and robust to IV violations, was applied to confirm gene-trait causal relationships when P_Bonferroni_ <0.05^99^. A merged reference panel of 10K UKBB and 1KG EUR (N = 4285)^100,101^ was used in GCTA^102^ to derive conditional/joint SNP groups.

### Cell Type Enrichment Tests

To evaluate whether identified cell-type-specific causal genes are overexpressed in corresponding GBM cell types beyond chance, we applied expression-weighted cell type enrichment (EWCE) v1.10.2^103^. Specificity values, indicating the proportion of a gene’s total expression in one cell type versus others, were calculated using snRNA for GBmap (core)^20^. We selected cell-type-specific gene groups with more than four genes, including significant and potential causal genes (passing association and replication studies), along with specificity matrices. After controlling for transcript length and GC-content, we executed EWCE with 10,000 bootstrap replicates, and applied Bonferroni correction to P-values.

### CT-FM-SNP

To pinpoint cell-type-specific causal variants in glioblastomagenesis, we implemented Cell Type Fine-Mapping SNP (CT-FM-SNP), which integrates GWAS fine-mapping data with cell-type-specific cCRE annotations (N = 927)^104^. The significant SNPs in the variants credible set underwent fine-mapping with S-LDSC results and were overlapped with candidate cis-regulatory elements (cCREs).

### Peak overlap

We overlapped the fine-mapping identified variants credible set in *EGFR* with astrocyte epigenetic data peaks. Bulk ATAC-seq^18^, snATAC-seq^17^, regulatory element data from ChIP-seq and PLAC-seq^17^ provided a comprehensive view of astrocyte-specific epigenetic effects. FORGEdb scores were retrieved via FORGEdb^29^.

### iCPAGdb

We used the interactive Cross-Phenotype Analysis of GWAS database (iCPAGdb) v1.1^105^ to identify glioma genetically shared traits across 3,793 phenotypes.

### Bidirectional MR-PheWAS

To determine genetic causal relationships, we performed bidirectional MR-PheWAS, including the largest available GWAS (before May 20, 2024) on genetic shared traits (P_Bonferroni_ <0.05). GWAS summaries from 208 traits were collected. Genome-wide significant (P <5×10^-8), independent (LD r² < 0.001, 10,000 kb) SNPs were chosen as instrument variables (IVs) via TwoSampleMR (v0.5.8)^106^. SNPs palindromic with intermediate allele frequencies or missing in the outcome GWAS were excluded. Radial MR (v1.1) detected and removed outlier SNPs^107^. We retained strong IVs (F>10) and phenotypes with >3 SNPs post-filtering to maintain statistical power. A Q-test P > 0.05 indicated no significant IV heterogeneity. MR-PRESSO (v1.0)^108^ evaluated horizontal pleiotropy; P>0.05 in global tests denoted no pleiotropic effects. The random-IVW method served as the main analysis given its higher power^109^, with P values Bonferroni-corrected. Weighted median (tolerating up to half pleiotropic SNPs)^109^ and 200-time simulation MR-cML-MA v0.0.0.9 (robust to correlated/uncorrelated pleiotropic effects)^110^ were used as sensitivity analyses. Reverse MR swapped exposure/outcome roles but applied the same filtering criteria; traits not significant (P_IVW_ > 0.05) were deemed non-bidirectional. Potential causal traits were required to (1) pass heterogeneity/pleiotropy checks, (2) lack bidirectional effects, and (3) be nominally significant (P<0.05) with consistent effect direction across all MR sensitivities.

### HDL

We estimated cross-phenotype genetic correlation between potential causal traits and glioma via High-Definition Likelihood (HDL) v1.4.0^111^, utilizing precomputed EUR LD matrices from UK Biobank. We focused on potential causal traits, P_HDL_ >0.05 indicated suggestive causal phenotypes, and genetically correlated (P_HDL_ <0.05) traits underwent further analysis.

### LCV

We employed the Latent Causal Variable (LCV) model^33^ on genetically correlated traits to filter false positives arising from genetic correlations. Traits with P < 0.05 and the same effect direction as MR analyses were considered suggestive causal.

### Gene-based PheWAS

We performed gene-based PheWAS analyses using GWAS ATLAS (3,302 unique traits from 4,155 GWASs)^112^ and UK Biobank TOPMed-imputed PheWeb (1,419 unique PheWAS codes from 400,000 EUR individuals)^113^ to annotate genetic pleiotropy. Genome-wide significant traits (P <5×10^-8) were reported.

### Drug Repurposing Analysis

We queried predicted drug-gene interactions from the Drug-Gene Interaction database (DGIdb v5.0)^114^, retaining only those with high interaction scores (> 0.8). DrugBank v6.0^115^, canSAR.ai v1.6.7^116^, Chinese Clinical Trial Registry (ChiCTR, https://www.chictr.org.cn/), ClinicalTrials.gov (https://www.clinicaltrials.gov/), and the WHO (https://www.who.int/) supplied information on chemical structure, late-stage clinical trials (pre–2024 July 20), and disease indications for drug repositioning.

Additional details are provided in the **Supplementary Methods**.

## Ethics

Ethical approval for this study was not required as our analyses were based on summary statistics from published GWAS or the data were publicly accessible.

## Acknowledgments

We thank the consortium studies such as GLIOGENE, AGOG, UKBB, FinnGen, and others. Their collaborative efforts and dedication were instrumental in enabling this study. We also appreciate Dr. Beatrice S Melin and Dr. Karen Alpen for their assistance in accessing the GWAS datasets.

## Author contributions

H.Y.F. and H.K.L. designed the analysis. H.Y.F. analyzed the data. H.Y.F. wrote the paper. H.Y.F., and H.K.L. provided feedback and approved the final version of the paper.

## Data available

The GLIOGENE consortium glioma GWAS summary statistics are available from the European Genome-phenome Archive (EGA, http://www.ebi.ac.uk/ega/) under accession number EGAS00001003372. AGOG glioma GWAS summary statistics are applied from AGOG. The FinnGen R9 glioma GWAS summary statistics are available from https://www.finngen.fi/en/access_results. Summary QTL database reference panel weights generated for this study used in TWAS and PWAS are available in https://figshare.com/articles/dataset/_b_TWAS_PWAS_gene_expression_prediction_models_b_/28080035. The reference panel, which is the merged genotype data from UK10K and 1000 Genomes Project Phase 3, is available from the EGA under accession number EGAD00001000776. The PLAC-seq, snATAC-seq and ChIP-seq data were downloaded from https://github.com/nottalexi/brain-cell-type-peak-files. pcHi-C was downloaded from GSE113481, bulk ATAC was downloaded from GSE113480. The remaining data are available within the Article, Supplementary Information or Source Data files.

## Code availability

We used publicly available software for the analyses. The software used is listed in the Methods section and supplement table. Custom analysis scripts are available at github (https://github.com/1667857557/Glioma-Cell-Type-Specific-Causal-Genes).

