## Supplementary material for "Single-cell multi-omic integration analysis prioritizes druggable genes and reveals cell-type-specific causal effects in glioblastomagenesis": Supplementary Figure.docx

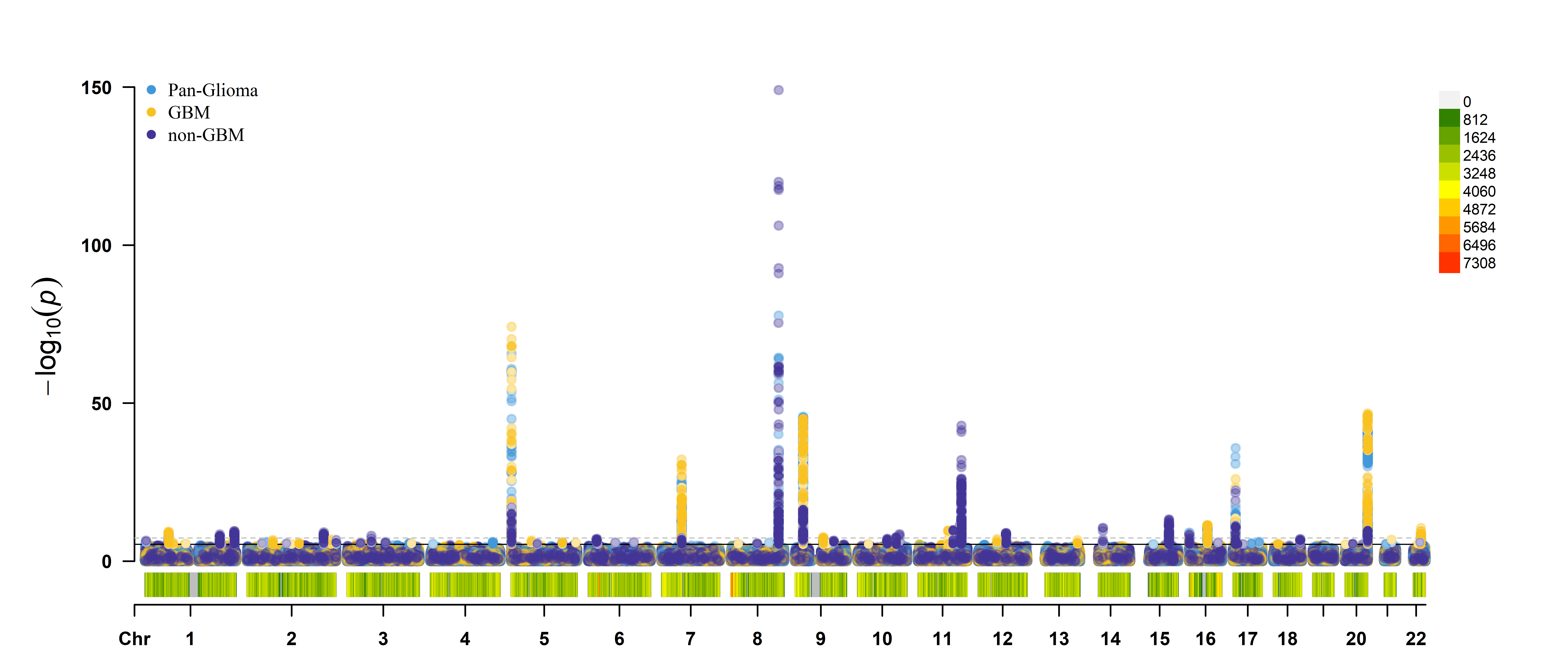

**Supplementary Figure 1**: Multiple traits GWAS Manhattan plot including pan-glioma, GBM, and non-GBM

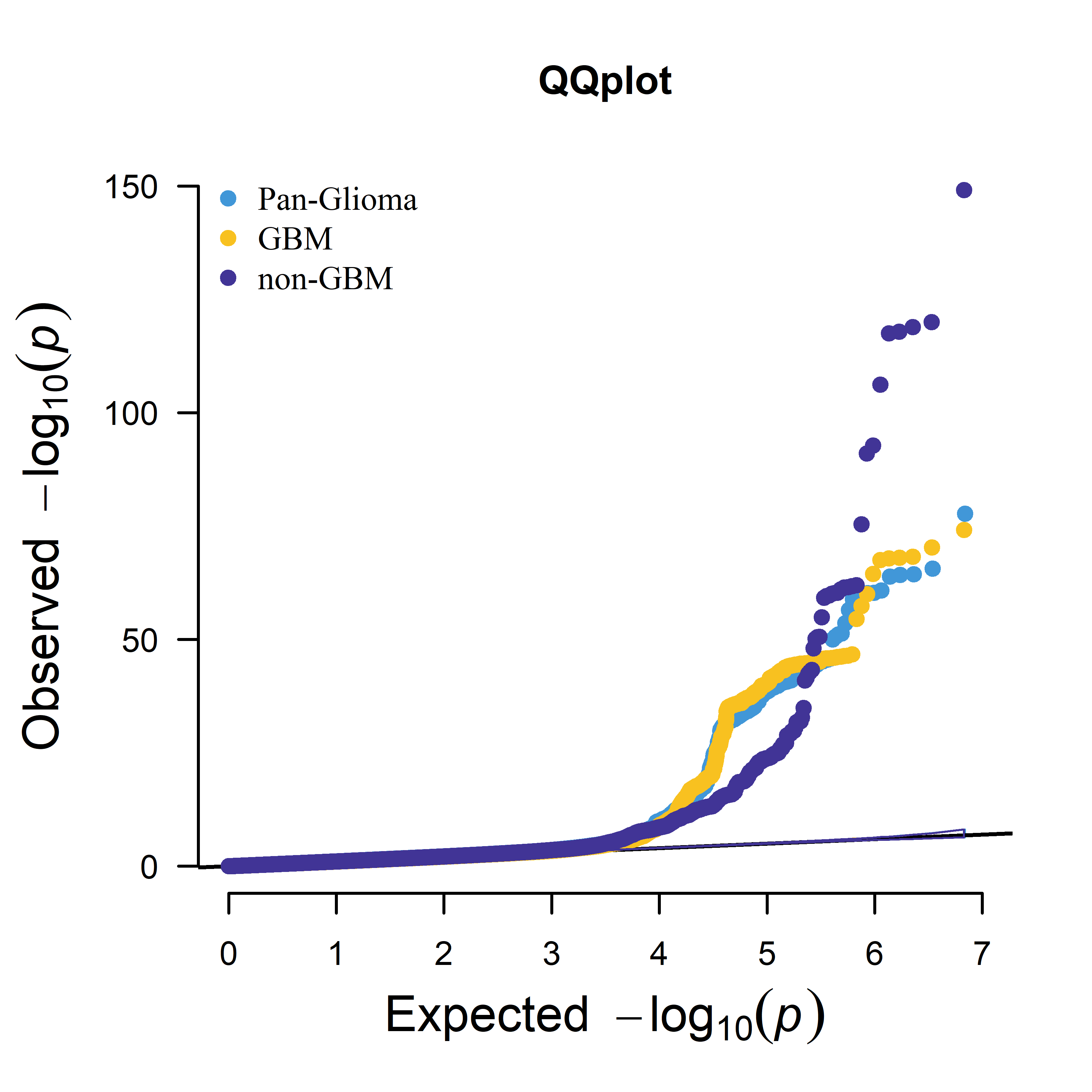

**Supplementary Figure 2**: Multiple traits GWAS QQ plot including pan-glioma, GBM, and non-GBM

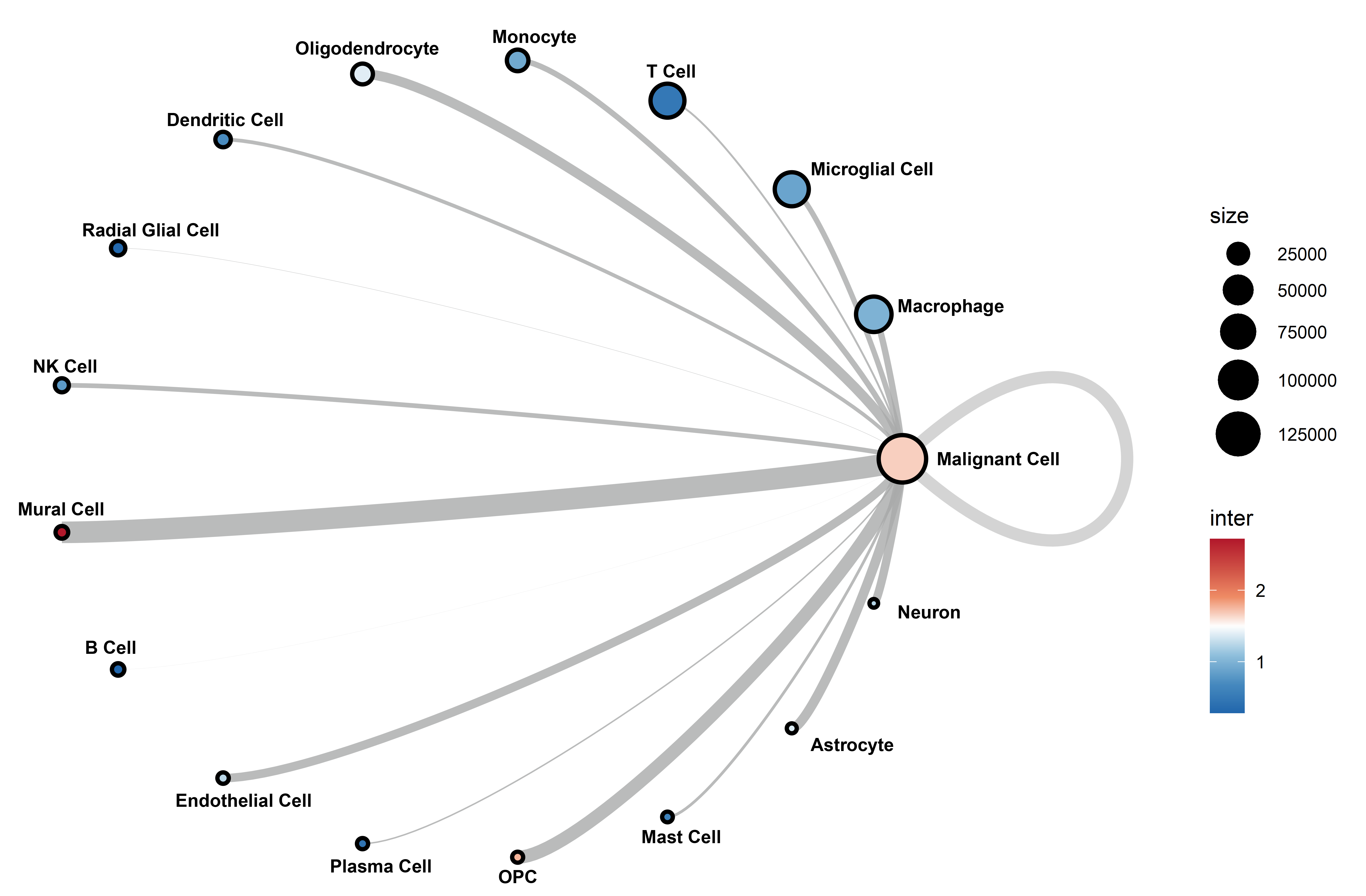

**Supplementary Figure 3**: Circle plot depicting cell-to-cell communication in glioblastoma (GBM) high-incidence brain regions, analyzed using the GBmap (expanded) single-nucleus RNA (snRNA) dataset.

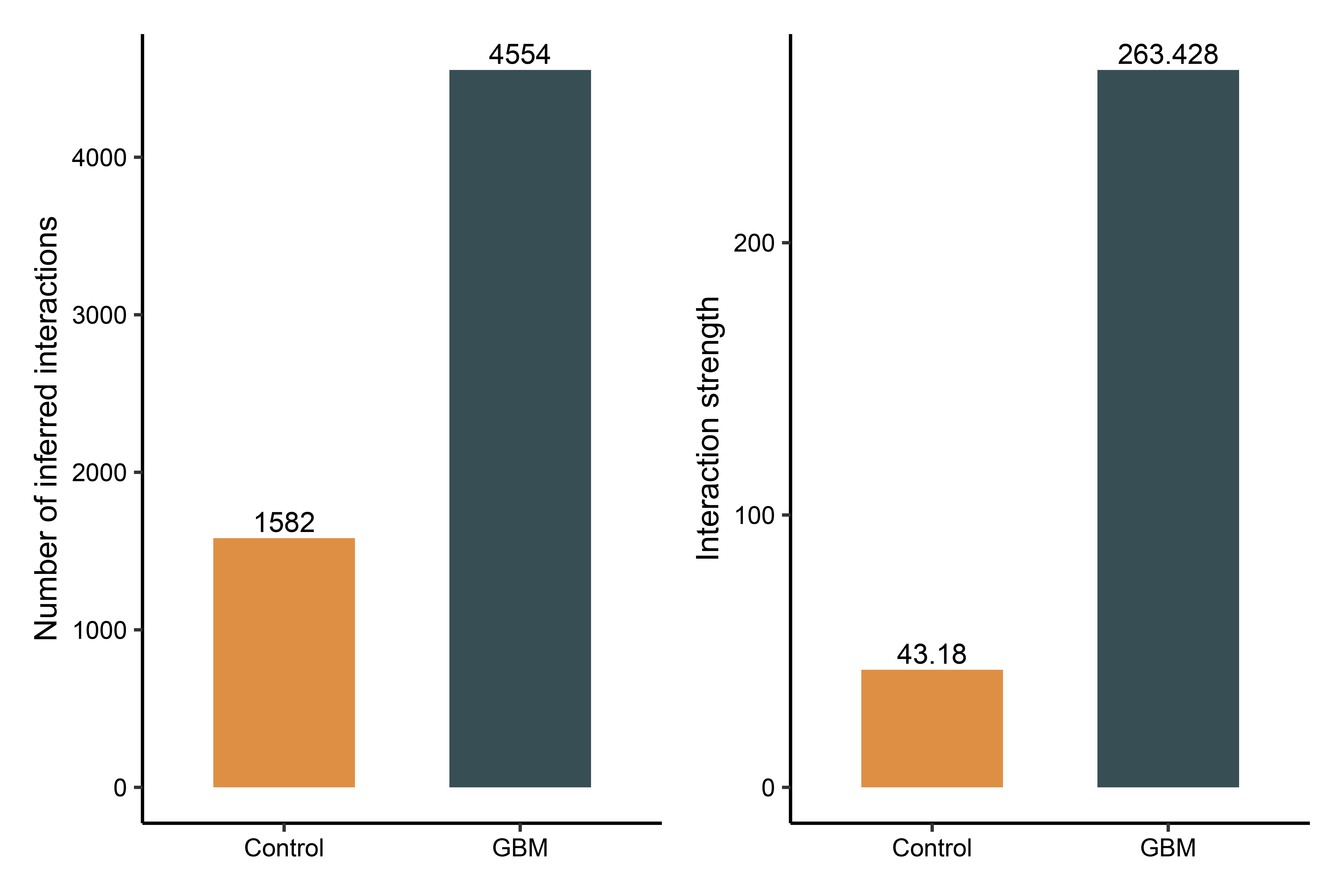

**Supplementary Figure 4**: Bar plots comparing the number (left) and strength (right) of inferred interactions between non-malignant cells in GBM high-incidence brain regions and healthy controls.

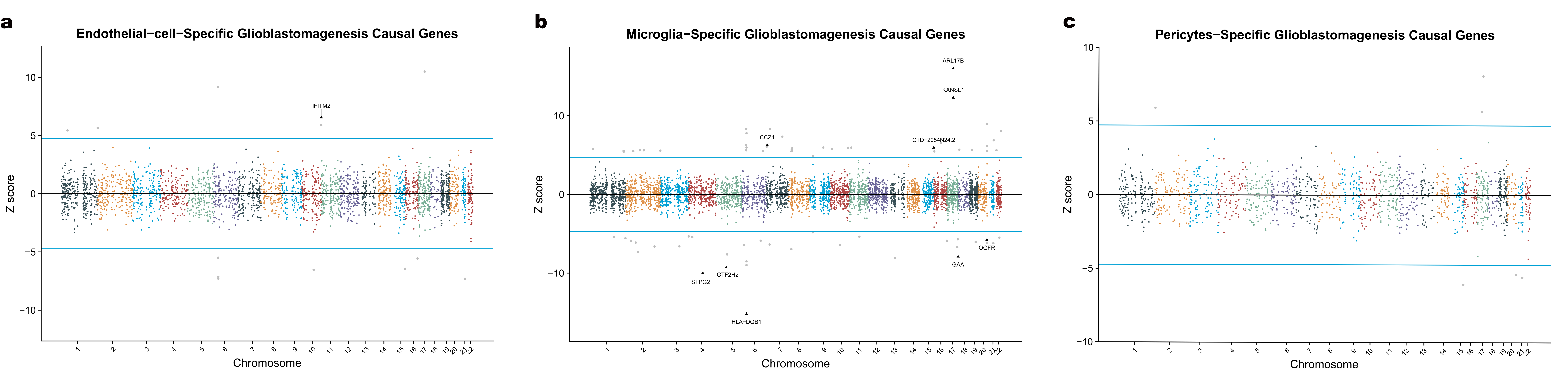

**Supplementary Figure 5**: **(a-c)** Manhattan-style plots displaying cell-type-specific glioblastomagenesis causal genes across chromosomes for Endothelial cell, Microglia, and pericyte. Z-scores represent gene effect size and direction in QTL-based association study, with significant cell-type-specific causal genes highlighted as red triangles (Duplicated in QTL-based association study, PP.H4.abf ≥ 70%, and with causal significance) and potential causal genes as black triangles (Duplicated in QTL- based association study, and PP.H4.abf < 70%). Grey dots indicate unduplicated genes from QTL-based association studies. Horizontal blue lines denote significant thresholds (P= 2.78e-6; 0.05/17,964).

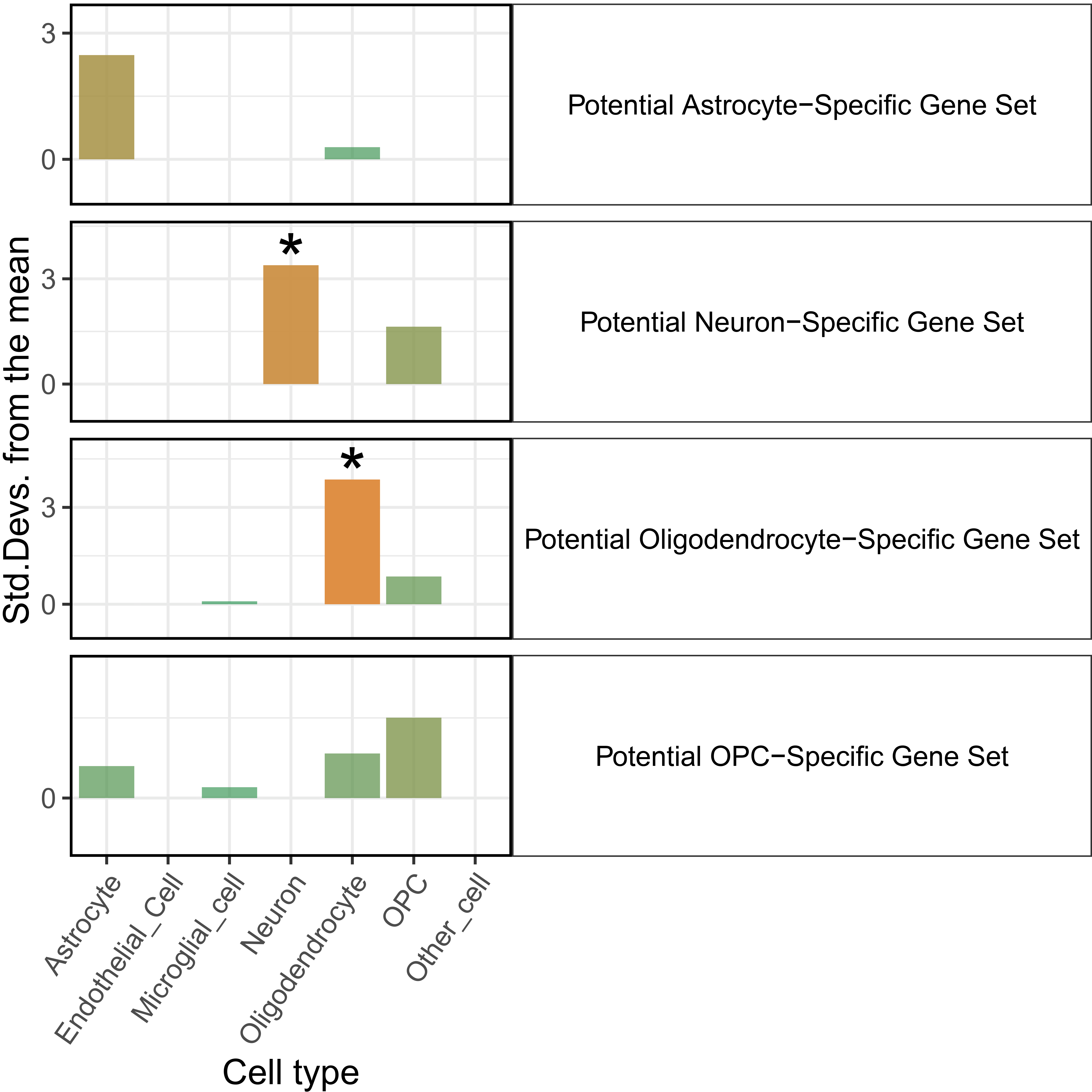

**Supplementary Figure 6**: Bar plots showing the cell type enrichment test in four potential cell-type-specific causal gene sets (astrocyte, neuron, oligodendrocyte, OPC). Asterisks (*) highlight significant deviations, indicating overexpression in cell-type compared to the bootstrapped mean.

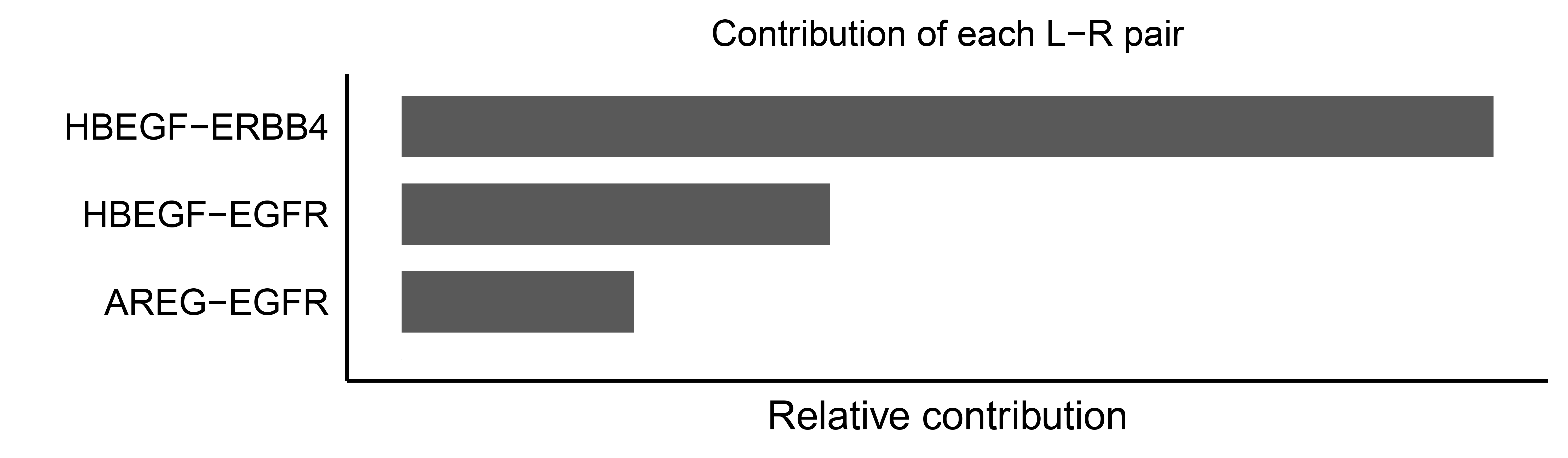

**Supplementary Figure 7**: Bar plot showing the main L-R pair and corresponding relative contribution in EGF pathway in GBM non-malignant cell data for GBmap (expanded).

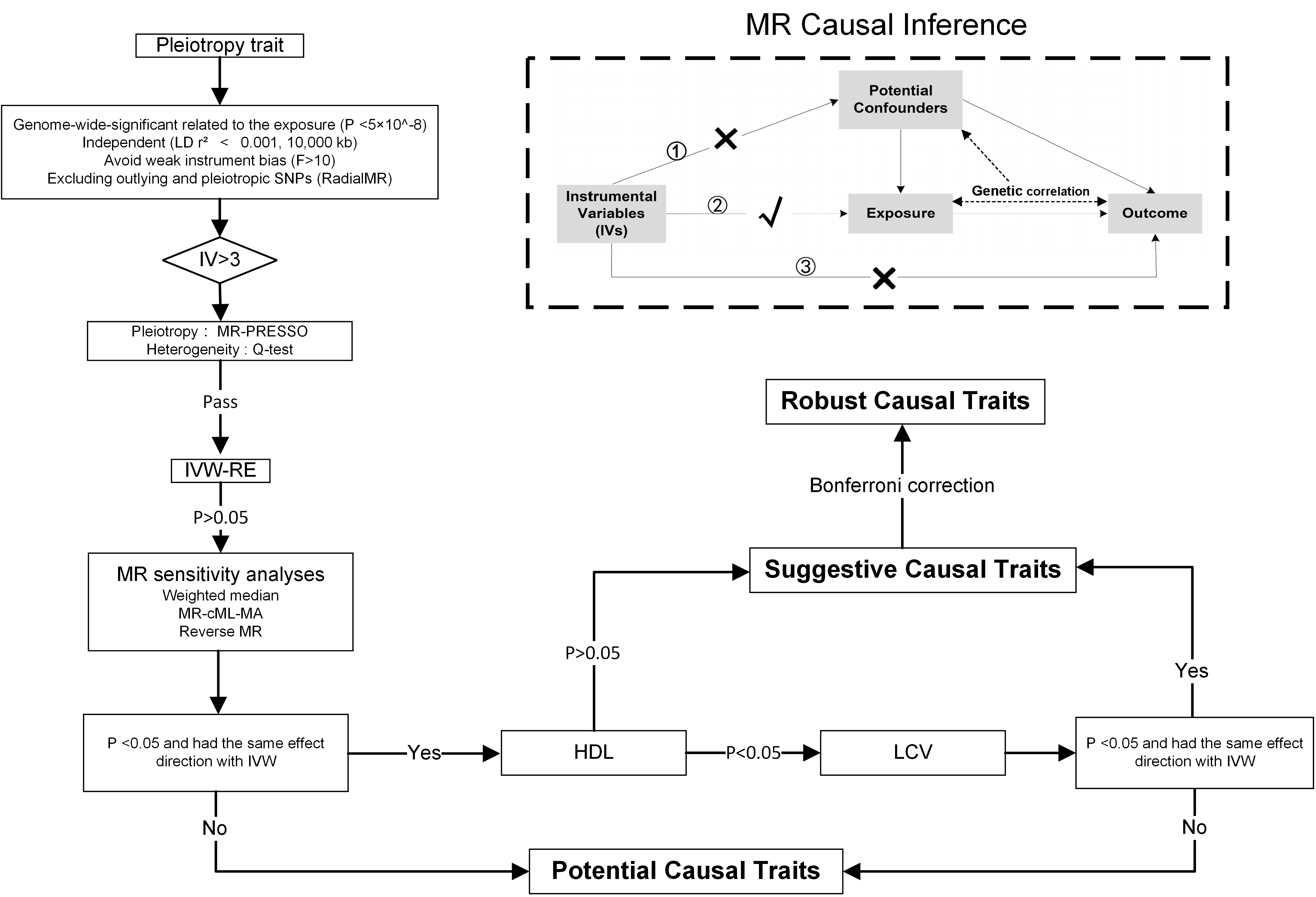

**Supplementary Figure 8:** Workflow of MR-PheWAS. When exposure and outcome are used for causal estimates in MR inference, three assumptions must be satisfied. ① Relevance assumption: the genetic variations are highly related to the exposure, ② Independence assumption: the genetic variants are not associated with any putative confounder of the association between exposure and outcome, and ③ Exclusion restriction: the variants do not alter the outcome independently of exposure.
