## Supplementary material for "Single-cell multi-omic integration analysis prioritizes druggable genes and reveals cell-type-specific causal effects in glioblastomagenesis": Supplementary Method.docx

**eQTL Data**

Brain-specific eQTL datasets were selected from whole brain (Nef  = 3,154)^1^, five different brain regions (Nef  = 108 to 2,683)^2^, and eight main brain cell types from two studies: one by Fujita et al.^3^, involving 1,509,626 cell nuclei from 424 EUR population individuals, and another by Bryois et al.^4^, involving 577,115 cell nuclei from 192 EUR population individuals. We focused on cis-eQTLs within ±1 Mb of the gene for all datasets. Detailed information on these studies is provided in **Supplementary Table 2**.

**pQTL Data**

Four brain proteomic studies from the EUR population were included: Wingo et al.^5^, with 10,198 proteins and Nef = 716; The Religious Orders Study and Rush Memory and Aging Project (ROS/MAP) cohort^6^, with 8,356 proteins and Nef = 376; The Banner Sun Health Research Institute (Banner) cohort^6^, with 11,518 proteins and Nef = 152; and Yang et al.^7^, with 1,079 proteins and Nef = 380. We focused on cis-pQTLs within ±500 kb of the gene across all studies. Detailed information on these studies is provided in **Supplementary Table 3**.

**sc/snRNA-seq data**

Three large-scale sc/snRNA-seq datasets were included in biological pathway enrichment analyses: (1) Pan-glioma scRNA-seq data generated by Johnson et al., containing 55,284 single-cell transcriptomes from 11 adult patients^8^ ; (2) GBM snRNA-seq data from GBmap (core), encompassing 338,564 cells from 110 IDH-wild-type GBM patients^9^; and (3) Non-GBM scRNA-seq data were generated by Wei et al.^10^, including 76,639 cells from 16 IDH-mutant glioma patients. Two large-scale snRNA-seq datasets from brain regions with high GBM incidence (frontal, temporal, and parietal lobes) were included in cell communication analysis: (1) GBM snRNA-seq data from GBmap (expanded), containing 289,395 single-nucleus transcriptomes from 240 adult patients^9^; and (2) healthy snRNA-seq data from the Human Brain Cell Atlas, containing 839,975 single-nucleus transcriptomes from 3 adults^11^. Detailed information on these studies is provided in **Supplementary Table 4**.

**Data Cleaning**

All GWAS and QTL summary data were standardized using MungeSumstats^12^. Briefly, chromosome positions in GRCh38 were lifted over to GRCh37 using the UCSC Chain file, duplicate variants and those with MAF < 0.01 were removed, and SNP IDs were mapped to dbSNP 155. For the sc/snRNA-seq data, we used the Seurat package^13^ to normalize and scale the data. The cell type annotations provided by the original studies were used.

**Gene prioritization using similarity-based methods**

**(1) PoPS**

Polygenic Priority Score (PoPS) identifies causal genes by scoring 18,383 protein-coding genes^14^. First, we conducted genome-wide gene-based association analysis (GWGAS) of 18,879 protein-coding genes using MAGMA^15^, with LD estimated from the 1000 Genomes Project Phase 3 EUR population^16^. The MHC region (chr6:28,477,797–33,448,354) was excluded before analysis due to its complex LD structure. We then computed PoPS for each gene by fitting a joint model for enrichment of features. Considering that the power of PoPS depends on the features included, we conducted analyses separately including all tissues and brain-specific features. Based on these scores, we ranked the genes within the genome-wide significant LD blocks, which contain more than one genome-wide significant SNP (5×10^-8) in each defining LD block^17^, and reported the top 500 (<3%) genes with the highest PoPS scores across all blocks as potential causal genes.

**Gene prioritization using locus-based methods**

**(1)** **Mapgen**

We ran fine-mapping using the Sum of Single Effects (SuSiE) method on GWAS summary statistics in the mapgen^18^. We first partitioned the genotype into 1703 LD blocks using LDetect^17^ and ran fine-mapping^19^ on LD blocks that contained more than one genome-wide significant SNP. This function inputs the SNPs' z-scores and pairwise LD matrix in a block. We used precalculated LD matrices, which were computed on 10% of the UK Biobank independent and White British samples (>50,000) for variants with MAF > 1%. The kriging_rss () function in SuSiE was used to detect and flip mismatched SNPs to avoid the effects of out-of-sample genotype information. We ran SuSiE with L = 10, which allows at most 10 causal SNPs for each LD block^19^. The posterior inclusion probability (PIP) of a gene is a weighted sum of PIPs of all SNPs linked to the gene. To improve long-range regulation causal gene discovery, we incorporated ABC scores generated from brain-related cell-types (H1_Derived_Neuronal_Progenitor_Cultured_Cells-Roadmap, bipolar_neuron_from_iPSC-ENCODE, and astrocyte-ENCODE)^20^, as well as brain chromatin loop data from snATAC-seq (neurons, microglia, oligodendrocytes, and astrocytes)^21^ and pcHi-C (excitatory neurons, hippocampal dentate gyrus-like neurons, lower motor neurons, and fetal astrocytes)^22^. We ranked the genes in genome-wide significant LD blocks; the gene with the highest PIP in each block and PIP > 0.8 was considered a potential causal gene.

**(2) TWAS**

We performed a transcriptome-wide association study (TWAS) using FUSION^23^, which estimates phenotype-related genes using GWAS summary data and precalculated gene expression prediction models^24^. We used the method described by Pain et al.^25^ to calculate the gene expression prediction models for QTL summary data when individual-level data were not available. Briefly, this method calculates ten TWAS prediction models using cis-QTL summary data and a reference panel from the 1000 Genomes Project Phase 3 EUR population, using eight leading summary statistic methods, We used a robust version of GCTA-GREML^24,26^ to estimate the heritability of gene expression, which generates heritability estimates per feature as well as the likelihood ratio test P value. Only features that have a heritability of P<0.05 were retained for TWAS analysis. Then, the TWAS-Fusion was used to estimate the TWAS association statistics between predicted gene expression and glioma by integrating information from expression reference panels (SNP-expression weights), GWAS summary statistics (SNP-glioma effect estimates) and linkage disequilibrium reference panels (SNP correlation matrix). Finally, the ACAT-O^27^ was used to combine the results of all predictive models, generating meta-P values and effect sizes^28^. SNPs in the MHC region were excluded from analyses due to their complex LD structure.

**(3) PWAS**

We performed a proteome-wide association study (PWAS) based on four different brain-specific cis-pQTL datasets measured by different proteomics techniques. Individual-level expression weights were generated using the ROS/MAP and Banner datasets^29^, and summary-level expression weights were generated by following the workflow described in TWAS^25^. LD information from the 1000 Genomes Project Phase 3 EUR population was used.

**(4) SMR**

Mendelian randomization (MR) provides causal inference based on Mendel's laws of inheritance; it was widely used in genetic drug target discovery^30^. We conducted summary-level Mendelian randomization (SMR) analyses^31^, which are based on a two-sample MR framework and allow summary-level GWAS input.

**(5) Replication studies**

To reduce false positive rates and enhance reproducibility in the QTL-based association analyses (SMR, PWAS, and TWAS), we conducted two separate replication studies using different QTLs and GWAS datasets. First, we conducted QTL-based association analyses using different source QTLs and the largest meta glioma GWAS dataset. Second, we included two independent European GWAS datasets: one from FinnGen R9 with a large sample size and standardized glioma classification, and another from AGOG with a reasonable case-control ratio (1:4) and available sex-stratified data, and previous QTL data to replicate the QTL-based association analyses. AGOG GWAS was used in the GBM replication study because most cases in AGOG were GBM^32^. Considering the imbalanced sample sizes between the discovery and replication studies, gene-disease pairs achieving the suggested significance thresholds (P_SMR_ <0.05 &P_HEIDI_ >0.05 or P_TWAS/PWAS_ <0.05) in two duplicate studies were considered successfully replicated and included in further analyses.

**(6) Colocalization**

We applied colocalization using the default priors in coloc and included the cis gene region. Colocalization analysis evaluates five hypotheses regarding the variant–trait associations: PPH0, no association with either trait; PPH1, association with expression of the gene, but not the glioma trait; PPH2, association with the glioma trait, but not expression of the gene; PPH3, association with the glioma trait and expression of the gene, with distinct causal variants; PPH4, association with the glioma trait and expression of the gene, with a shared causal variant.
